# Assessment of SARS-CoV2 genome sequence recovery from four lateral flow device products available in the UK

**DOI:** 10.1101/2025.01.23.25321032

**Authors:** Kuiama Lewandowski, Matthew Stokes, Alexandra Alexandridou, Abigail Fenwick, Mia L. White, Jack Crook, Karly-Rai Rogers-Broadway, Catherine Ryan, Deborah A. Williamson, Richard Vipond, Steven T. Pullan

## Abstract

Lateral flow tests have played a key role in the response to the COVID-19 pandemic and are likely to be a major component of diagnostic strategies to combat future outbreaks of infectious disease. One challenge posed by widescale use of lateral flow tests in the community is the loss of sequence information to track virus evolution and epidemiology.

Beyond their primary diagnostic function, it has been demonstrated that recovery of viral RNA for genome sequencing purposes is possible, from positive lateral flow devices (LFDs).

To assess the robustness and broader applicability of this process, we assessed SARS-CoV-2 RNA recovery and sequencing from the four major LFDs in use in the UK.

Testing both cultured virus and residual clinical nasal swab samples demonstrated that sequencing from LFD eluates is possible, at clinically relevant titres, within a reasonable processing time frame post-use, and gave sequences concordant with routine methods, but results varied across the four devices used.

This highlights the requirement for refinement of existing LFDs or of second–generation LFD design, where sequencing is an intended output from positive LFDs.

## Introduction

Genomic sequencing of SARS-CoV-2 has had a major role in the public health response to the COVID-19 pandemic, enabling mapping of viral transmission at global and local levels, informing infection control measures, and identifying and tracking the emergence of new variants (1).

Viral transport media from nasal/oral swabs have been the most widely utilised sample source for SARS-CoV-2 WGS pipelines. Typically, these have been residual material from diagnostic testing, taken forward for genome sequencing (most commonly via whole genome tiling amplicon-based methods) upon positivity (2). However, point-of-care lateral flow device (LFD) tests, including self-test kits are replacing PCR as diagnostics in many settings (3), meaning opportunities for genomic characterisation of circulating variants are increasingly limited.

Multiple studies have demonstrated the possibility of retrieving viral nucleic acid from positive LFDs. *Martin et al*., 2022 described an approach for whole-genome sequencing of SARS-CoV-2 from positive LFDs and demonstrated the application of this technique to PanBio and InnoScreen devices collected as part of clinical care. The same device brands were also used to demonstrate detection and whole genome sequencing of non-SARS-CoV-2 respiratory viruses present on the COVID-19 LFD (5). *Macori et al*., 2022 *d*emonstrated SARS-Cov-2 sequencing was possible from PanBio and Clinitest devices. *Rector et al*., 2022 demonstrated that genomic material of SARS-COV-2, influenza virus, adenovirus and rotavirus can also be retrieved from their corresponding positive LFDs, tested across 9 different device brands (Roche, FlowFlex, Newgene, Coris, Alltest, Boson, OrientGene, AMP and Nadal), showing stability for up to three months post-use storage at room temperature, but noting that there were significant differences in RNA yields obtained from different device buffers, highlighting the requirement for individual device testing prior to routine use in sequencing.

In this study we aim to perform an assessment of sequencing from the LFD, focussed on those LFD brands currently in use in the UK, with a focus on the feasibility of performing the analysis in real-world relevant processing time frame.

## Methods

### Lateral Flow Devices assessed

Four brands of SARS-COV-2 LFD widely used in the UK were procured for assessment for recovery of SARS-COV-2 genomic material :

- FlowFlex COVID-19 Antigen Rapid Test (FlowFlex)
- COVID-19 SureScreen Lateral Flow Self-Test (SureScreen)
- OrientGene Rapid Covid-19 (Antigen) Self-Test (OrientGene)
- INNOVA SARS-CoV-2 Rapid Antigen Test (Innova)

### Inactivated SARS-COV-2

Cell-cultured and X-ray-irradiation-inactivated SARS-COV-2 virions were provided by Virology and Pathogenesis group, UKHSA Porton Down. A dilution series of samples was produced ranging from 10^5^ to 10^1^ PFU/ml (RT-qPCR C_t_ vales of 18, 25, 28, 32 and 35).

### Clinical samples

Anonymous residual VTM from nasal swab samples was provided from routine UKHSA SAR-COV-2 surveillance sequencing.

### LFD loading

Mock or clinical samples were diluted 1:1 v/v with the corresponding LFD sample loading buffer and 70 µl applied to the device via pipette. After 15 min the LFD result was read. Ambient incubation was continued for 45 min before elution.

### Addition of preservatives

Where stated, 70µl of the tested preservative were added to the device via the sample loading port once the result had been read. Viral sample titre for all preservative testing 10^3^ PFU/ml (RT-qPCR C_t_ 25).

## LFD Elution

### Strip removal method

The LFD was opened manually along the join line of the outer casing, the internal strip was removed using two pairs of tweezers, placed into a 2 ml tube containing 700µl of MagMAX Viral/Pathogen Binding Solution (extraction buffer) and incubated for 60 min prior to nucleic acid extraction.

### Centrifugation method

700 µl of extraction buffer was added to the LFD via the sample loading port. The device was placed inside a 50 ml falcon tube, laid flat (face up) and incubated for 60 min. The tube containing the LFD was centrifuged at 2000 rpm for 2 min to elute the extraction buffer from the device into the falcon tube prior to nucleic acid extraction.

### Nucleic Acid extraction

Samples were extracted via the KingFisher Flex platform using the MagMAX Viral/Pathogen Nucleic Acid Isolation kits and 700µl of sample input, as per manufacturer’s instructions.

### SARS-COV-2 Sequencing and lineage calling

Samples were processed by the UKHSA Pathogen Genomics SARS-Cov-2 Sequencing Service, utilising the ARTIC network tiling amplicon scheme protocol (8), with primer scheme version 5.3.2. (<u>SARS-CoV-2 version 5.3.2 scheme release - Laboratory - ARTIC Real-time Genomic Surveillance</u>) and sequenced on an Oxford Nanopore GridION X5. Data analysis was performed by following the ARTIC network nanopore protocol for nCoV2019 (<u>10</u>) and lineage determined via Pangolin (11).

### RT-qPCR

RT-qPCR was performed using New England Biolabs, Luna SARS-CoV-2 RT-qPCR Multiplex Assay Kit. Assays were performed as per manufacturer’s instructions for the multiplex detection of 2019-nCoV_N1 and 2019-nCoV_N2 targets. Reported C_t_ values are for the N1 target.

## Results/Discussion

### Preliminary testing of recovery utilising inactivated SARS-COV-2

A titration range of inactivated SARS-COV-2 virions was used to assess the four device brands, with genomic material recovered by removal of the test strip (see methods).

We observed recovery of sufficient sequence information for lineage assignment (Typically > 70% of total genome) at both 10^5^ and 10^3^ PFU/ml concentrations (Equivalent to a sample C_t_ of 18 and 25 respectively) from three of the four devices tested (Figure 1). SureScreen being the exception, with reduced genome recovery across all concentrations tested. Samples with a concentration of 10^2^ and above consistently produced a positive LFD on all brands tested.

**Figure 1.**
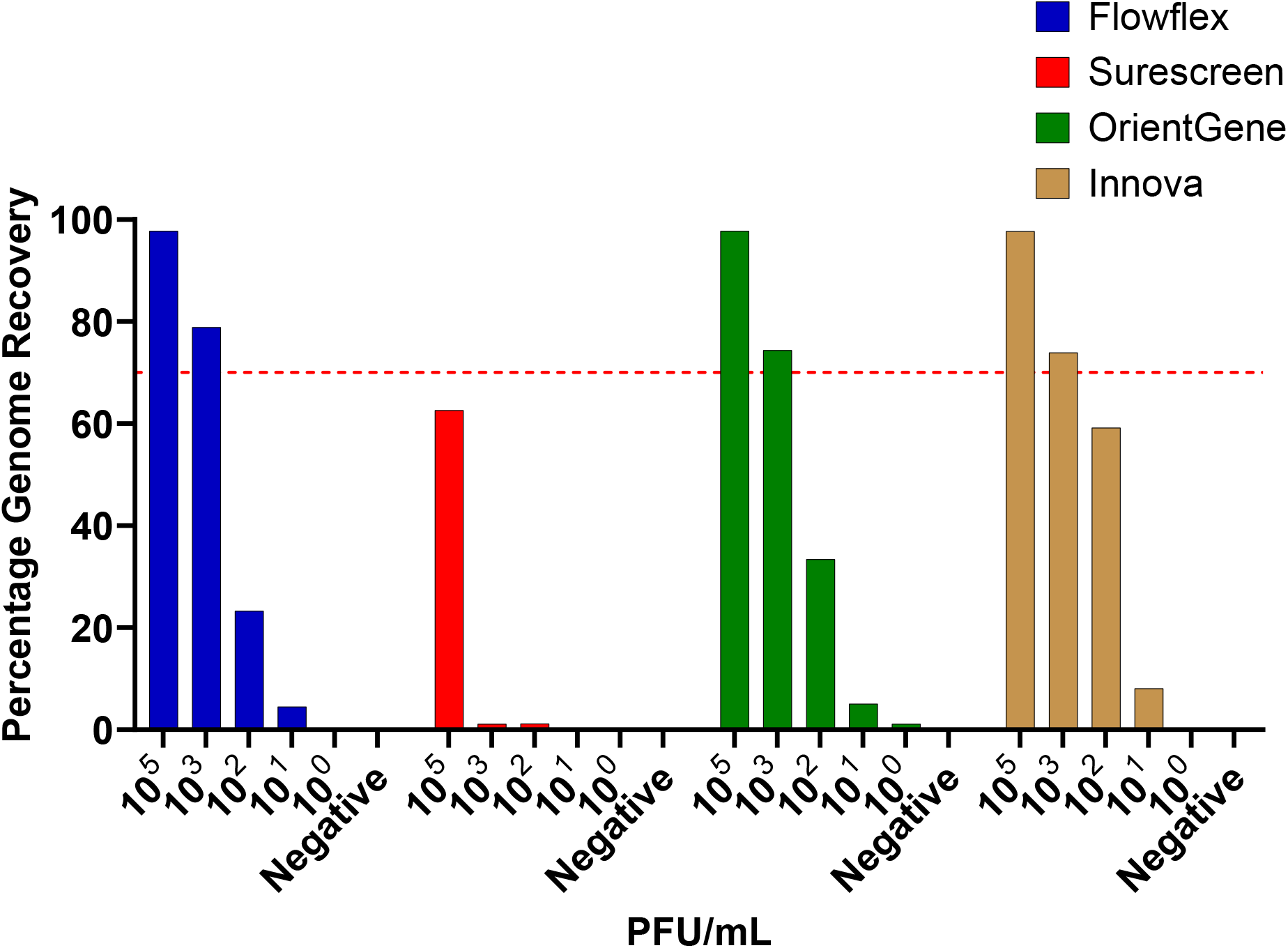
Percentage of total genome sequence recovered following elution of SARS-COV-2 virions from four brands of LFDs. Red line indicates 70% genome recovery (approximation of level required for lineage determination).

### Clinical sample testing

To assess recoverability from a clinically relevant sample type, we tested 40 residual nasal swab VTM samples (used in COVID-19 surveillance sequencing). Residual material from each sample was loaded upon each of the four LFD types for direct comparison.

Results again varied across devices. From OrientGene and Innova devices 80% of samples achieved sufficient coverage for variant calling, whereas for FlowFlex and SureScreen devices only 25 and 20% of samples produced sufficient genome coverage respectively (Figure 2).

**Figure 2.**
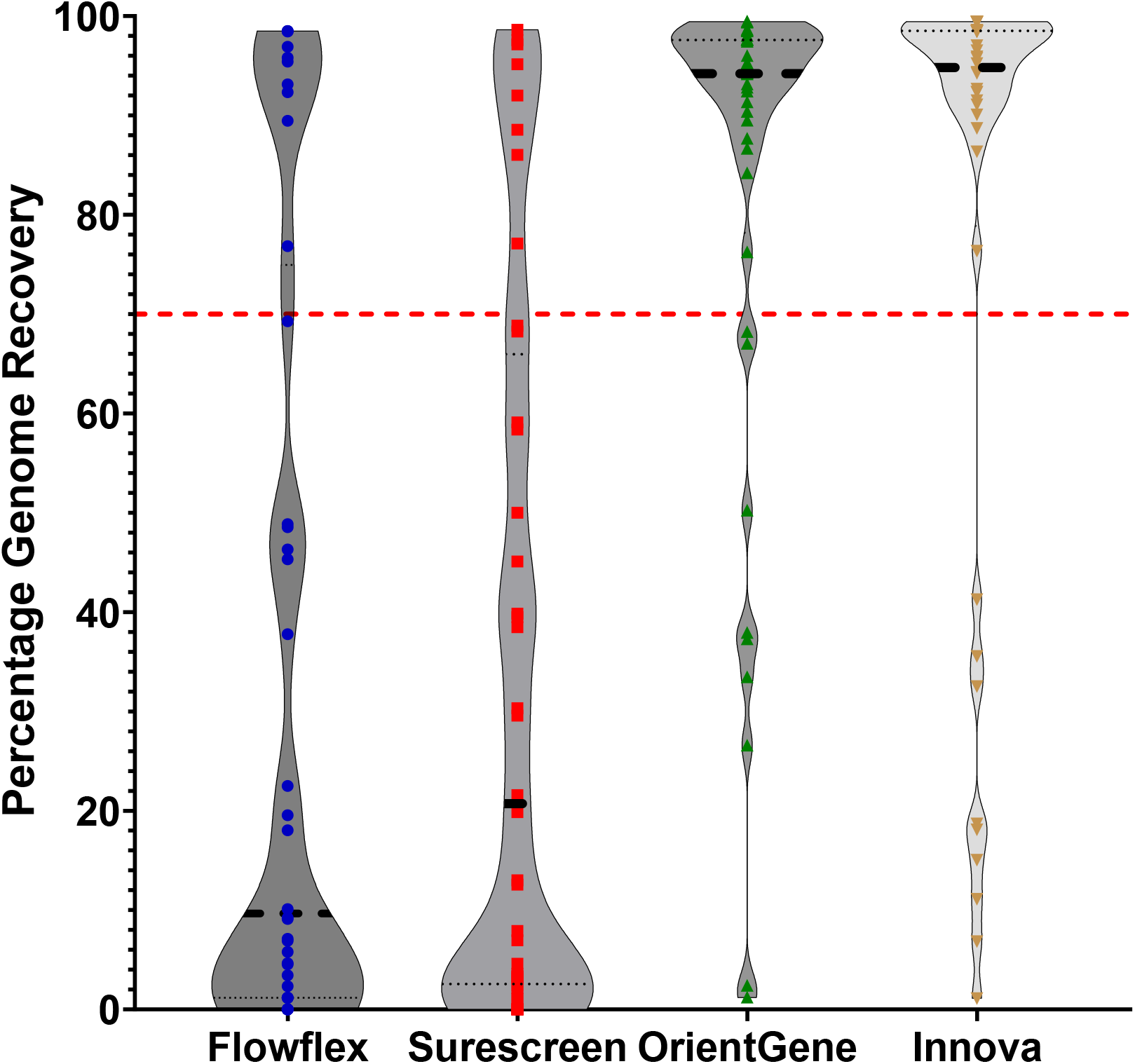
Percentage of total genome sequence recovered following elution of clinical samples across each of the four devices. Red line indicates 70% genome recovery.

In all cases where a lineage call was achieved, they were concordant with the expected result from initial sample sequencing.

### Elution efficiency and genome recovery

To assess efficiency of elution of viral nucleic acid from the LFDs, and the effect upon sequence recovery, RT-qPCR was performed on a subset of the clinical samples and corresponding LFD eluates. A similar level of reduction in viral concentration was seen between the primary sample and the LFD eluate for all devices (Figure 3) with mean C_t_ increases being 14.9 from FlowFlex, 13.3 from Surescreen, 11.4 from OrientGene and 12.5 from Innova.

**Figure 3.**
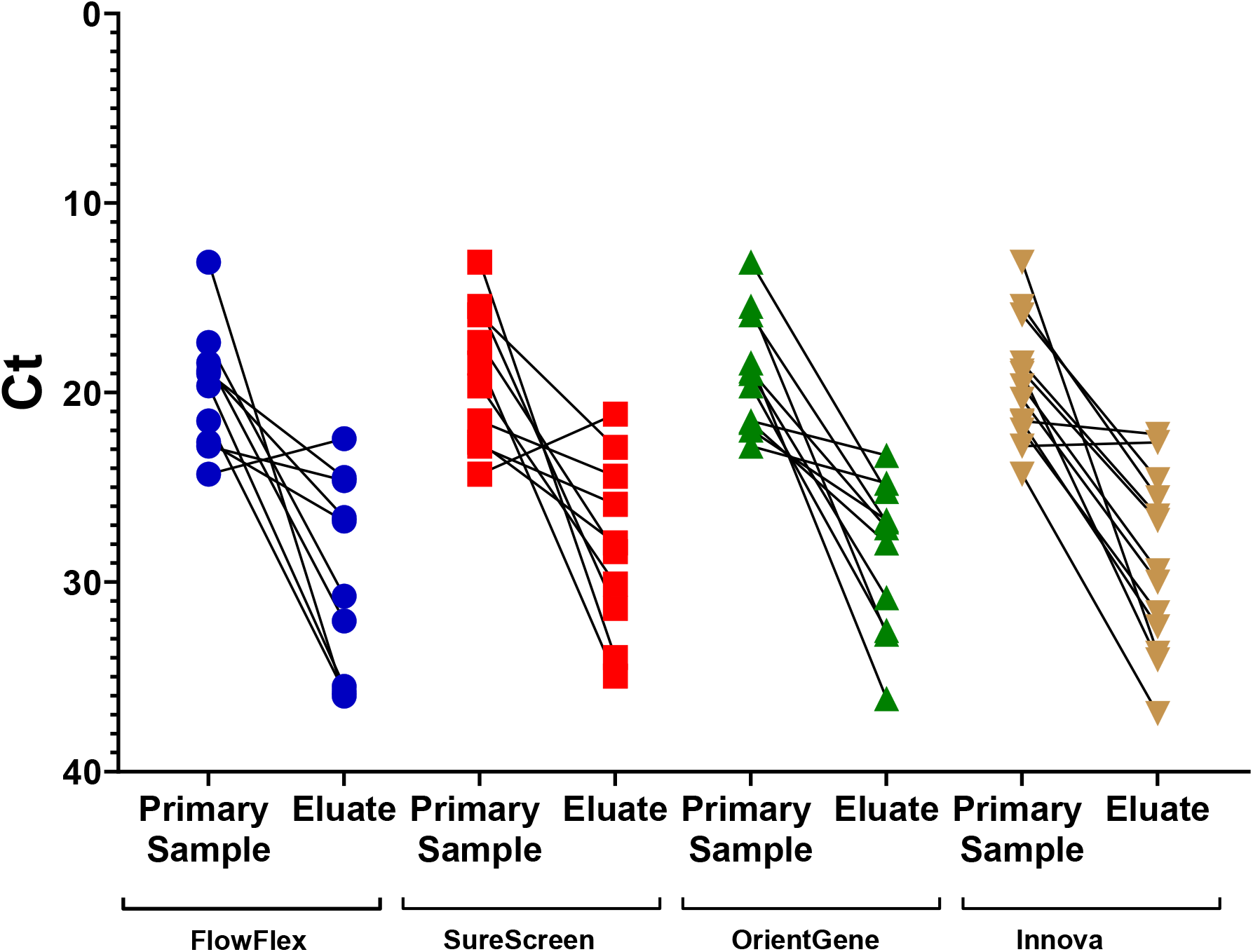
qRT-PCR C_t_ from original sample and in the corresponding material eluted from LFD.

No clear correlation was seen between the C_t_ of the eluted material and the percentage of genome recovered for FlowFlex and Surescreen devices. For OrientGene and Innova devices a trend was observed of C_t_ below 30 being sufficient to provide >70% genome recovery consistently, with higher C_t_ values providing more mixed results (Figure 4).

**Figure 4.**
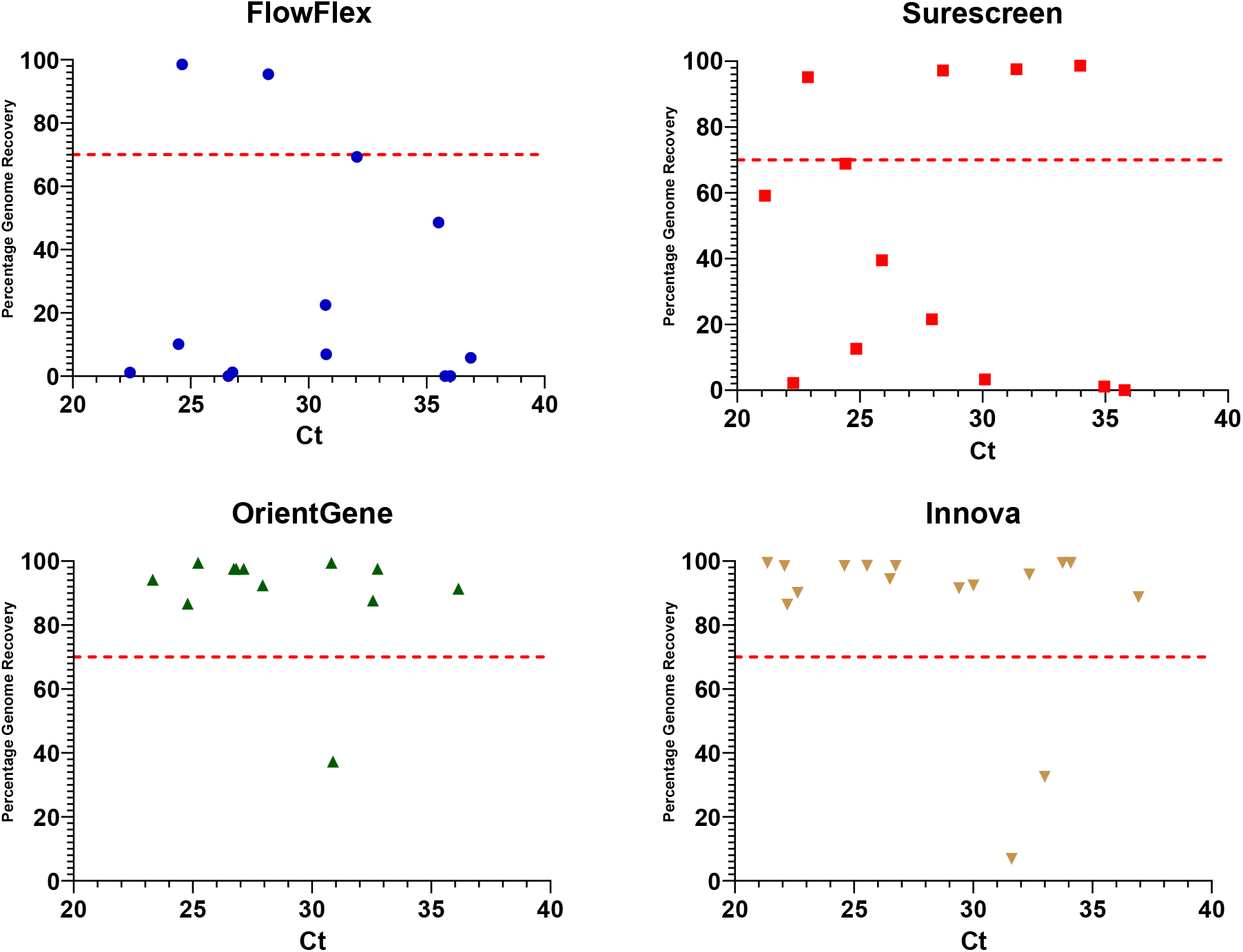
Comparison of percentage consensus genome sequence recovery from clinical samples compared to the viral titre of material eluted from the used LFDs. Red line indicates 70% genome recovery.

### Process optimisation and evaluation of the centrifugation method using clinical samples

Experiments initially followed the procedure for LFD strip extraction as set out by *Martin et al*., 2022. This process is time consuming and labour intensive, therefore, we set out to test the process of elution of sample in extraction buffer directly from the LFD via centrifugation as described by Macori *et al*., 2022.

For initial comparison the irradiated SARS-CoV-2 series, was processed in triplicate, across three time points, on each device by both methods. LFD centrifugation provided equivalent or improved genome coverage for all device brands (See Supplemental Table 1). A further comparison between the two extraction methods was made using residual clinical samples. The clinical samples tested were not paired but comprised of two batches containing samples with a clinically representative C_t_ range. Results from elution by centrifugation remained comparable to those from the strip removal extraction method when using clinical samples (Figure 5). The percentage of samples with a positive lineage call, comparing strip removal to centrifugation was 21.7% to 29.4%, 21.7% to 17.7%, 78.2% to 76.4% and 78.2% to 76.4%, for FlowFlex, SureScreen, OrientGene and Innova devices respectively.

**Figure 5.**
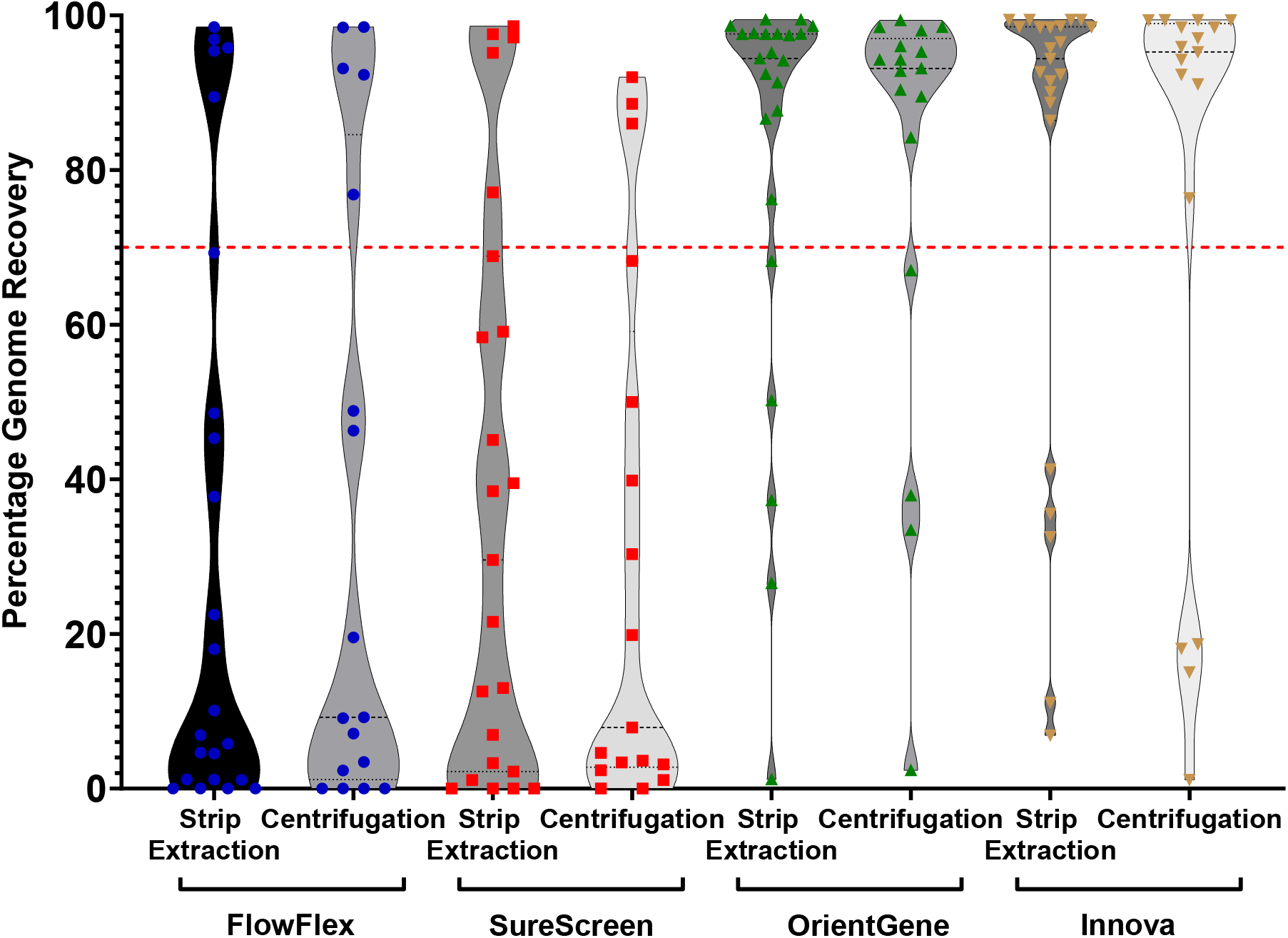
Distribution of percentage consensus genome sequence recovery following elution via strip removal (opened) or centrifugation (spun) method from clinical samples for all four LFD devices tested. Red line indicates 70% genome recovery.

### Assessment of sample viability upon LFDs over seven days of ambient incubation

Real-world use of LFDs as a sample source for sequencing applications raises logistical issues that are likely to include delays between initial sample LFD testing and laboratory processing, particularly in home-use scenarios. We set out to assess the impact of delayed time of LFD sample receipt and processing on the ability to recover informative genome sequences. An irradiated SARS-CoV-2 dilution series was used to assess the effect of such delays ranging up to seven days in duration. Stability of the nucleic acid in the used LFD as measured by genome sequence recovery was assessed for each device, from dilution in LFD buffer without being run on the device to running followed by multiple days incubation upon the device post-use, at room temperature, prior to extraction (Figure 6).

**Figure 6.**
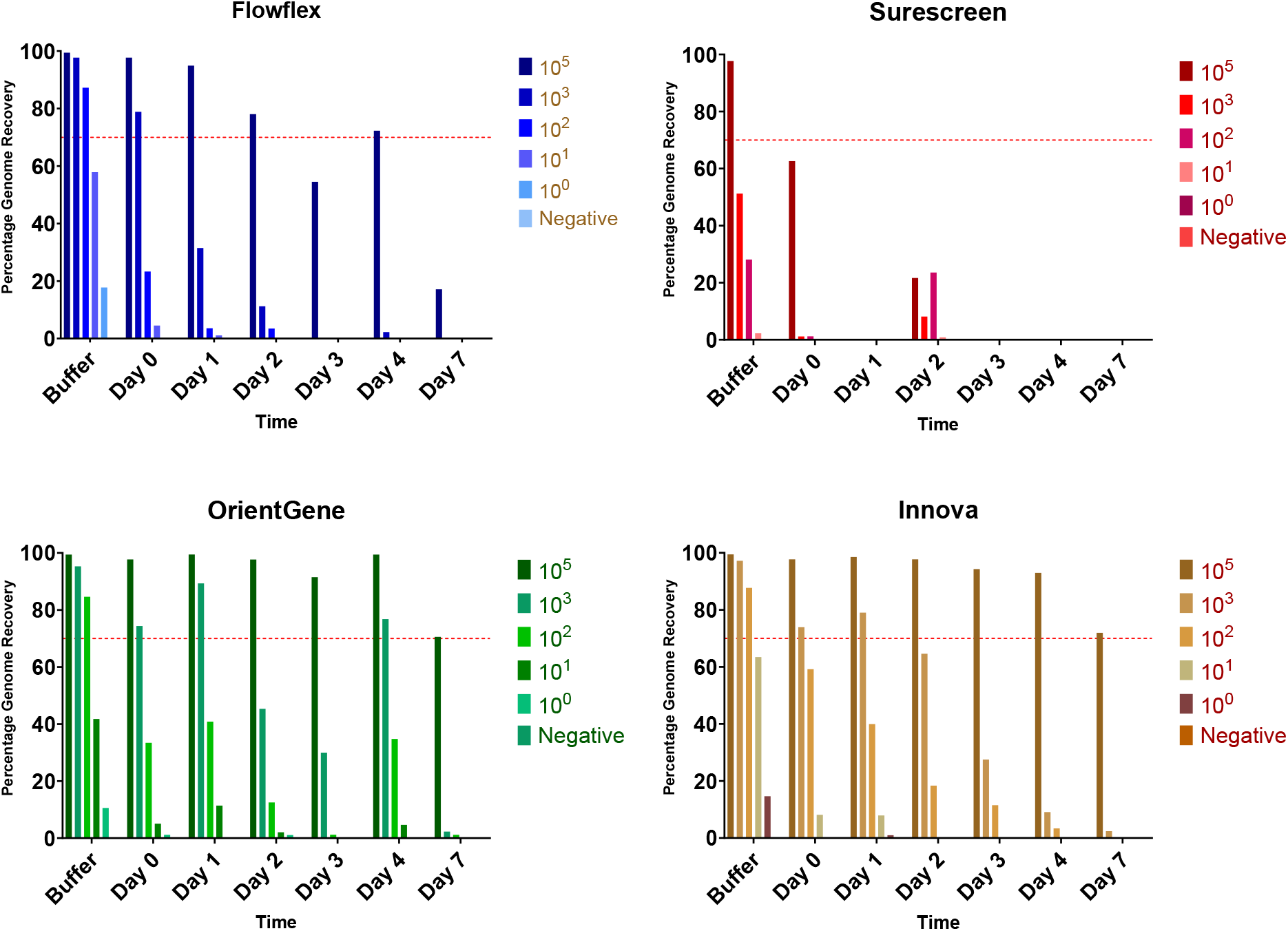
Percentage consensus genome sequence recovery following elution of a range of titres of SARS-COV-2 virions from the four LFD devices tested. Buffer indicates incubation in buffer without being run on a device. Red line indicates 70% genome recovery.

Decreased genome recovery was seen with increased delay time for all devices. The highest titre tested (equivalent C_t_ 18) provided sufficient genomic coverage for multiple days on FlowFlex, OrientGene and Innova devices (Figure 4) but the signal rapidly degraded on Surescreen devices. The C_t_ 25 equivalent sample showed inconsistent signal over multiple days on OrientGene and Innova devices but was rapidly degraded on FlowFlex and SureScreen devices.

### The effect of delayed time to elution on recovery from LFDs using residual clinical sample material

Residual clinical samples were used to assess the effect of delayed time to elution on recovery from LFDs. Ten clinical samples were tested on each of 4 LFD brands over the course of 3 days. Decreased genome recovery is seen over time in all cases, but OrientGene and Innova devices still provided sufficient genome coverage for lineage calling from 60% and 50% of clinical samples tested after 2 days incubation. FlowFlex and SureScreen produced reduced genome coverage from the samples tested, even in the absence of delayed recovery (Table 1).

**Table 1.** Percentage of clinical samples from which sufficient genome sequence was recovered from each LFD brand following either immediate extraction for sequencing (Day 0) or ambient incubation on the device (Day 1, Day 2).

|  | Percent of samples with Lineage Call |  |  |
| --- | --- | --- | --- |
|  | Day 0 | Day 1 | Day 2 |
| FlowFlex | 30 | 20 | 0 |
| SureScreen | 30 | 20 | 0 |
| OrientGene | 90 | 60 | 60 |
| Innova | 80 | 70 | 50 |

### The effect of nucleic acid preservatives on recovery from LFDs over five days of ambient incubation

Addition of three RNA stabilising agents to devices post running of a mid-titre SARS-COV-2 mock sample (10^3^ PFU.ml) show varied results. DNA/RNA shield and RNALater preserved RNA and allowed near complete genome sequencing after 5 days room temperature incubation on both OrientGene and Innova devices. Inhibisure was similarly effective on the Innova device but showed no preservative effect on OrientGene devices. Results on FlowFlex were inconsistent for all preservatives tested (Figure 7).

**Figure 7.**
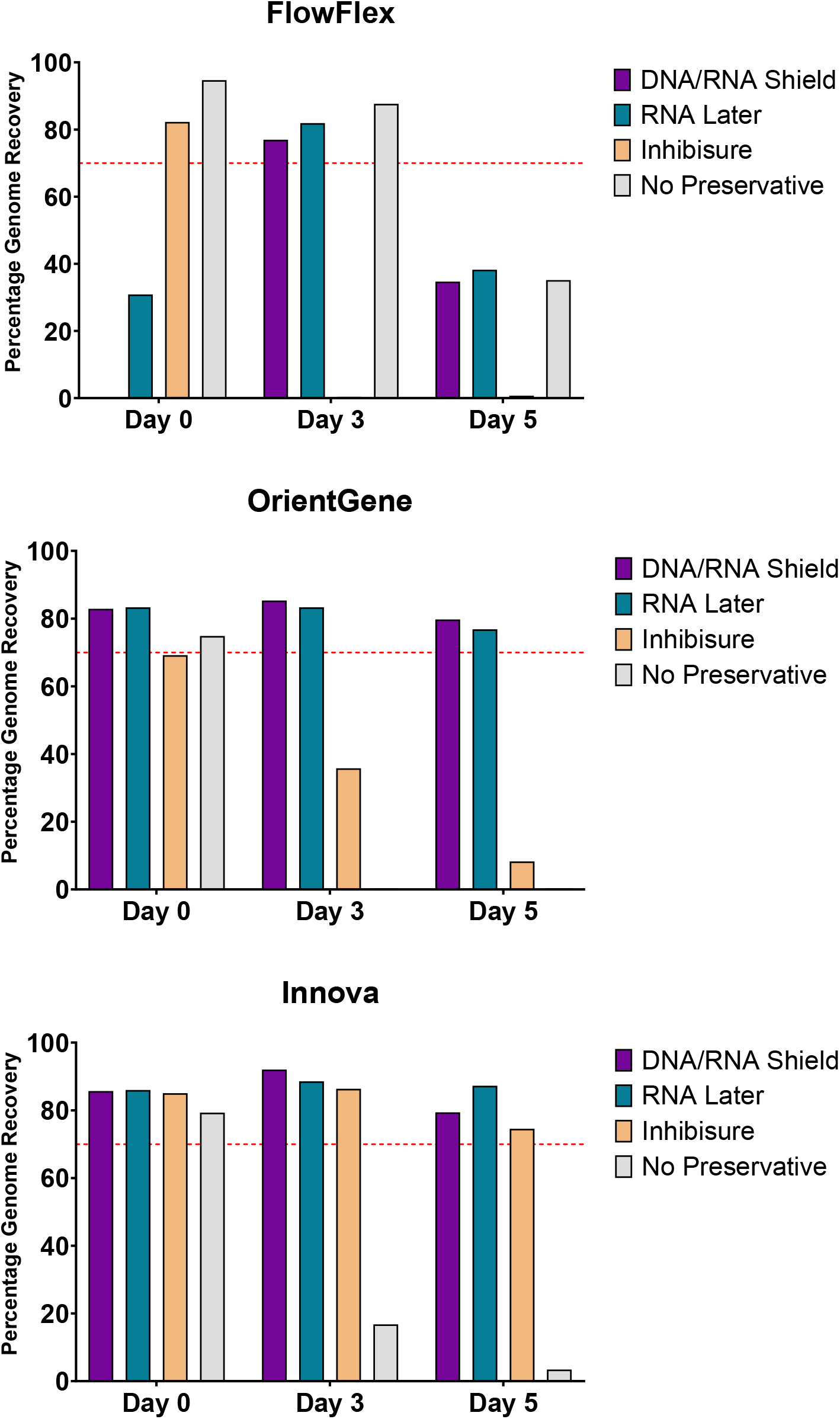
The effect of nucleic acid preservatives on genome recovery from LFDs over five days of ambient incubation. Red line indicates 70% genome recovery.

## Discussion

In this study we set out to assess the feasibility of SARS-CoV-2 amplicon sequencing using LFD eluates from four major LFD brands currently in use in the UK, with the aim of generating a protocol that could subsequently be expanded to other potential use settings, such as support for future variant monitoring programs. The LFDs used in this study were not designed to be compatible for use with downstream genomic studies, as such device performances are not a reflection of their ability to detect SARS-CoV-2 when used for their intended purpose.

Our evaluation using inactivated SARS-COV-2 observed recovery of sufficient sequence information for lineage assignment at titres as low as 10^3^ PFU/ml (sample Ct ∼25) from three of the four evaluated major devices currently in use within the UK.

Our assessment of genome recovery from 40 residual clinical nasal swab samples (C_t_ range from 13 to 24) for each LFD type found reduced genome recovery for clinical samples from FlowFlex and SureScreen devices (25% and 20% of samples with a lineage call respectively) when compared to OrientGene and Innova devices (80% and 80% of samples with a lineage call respectively).

Rector *et al*., 2022 tested a range of LFD brands, using stored nasopharyngeal samples, they found that LFD buffer components can be deleterious for the viral genetic material, resulting in lower RNA concentrations and interfering with sequencing. In our study, a similar reduction in viral concentration was seen in LFD eluates of all brands compared to the originating clinical samples. Indicating that reduced elution efficiency is unlikely to be the cause of poorer genome recovery via amplicon sequencing in some devices. Increased levels of fragmentation or the presence of inhibitors may be significant factors.

The process of removing strips from LFD casings for nucleic acid extraction is time consuming, labour intensive and not feasible for use in high throughput processing. Work by Macori *et al*., 2022 described an extraction method by centrifugation. In our study, we confirmed LFD centrifugation provided equivalent or improved genome coverage for all device brands.

Decreased genome recovery was seen with increased delay time for all devices. OrientGene and Innova showed the best recovery over time, with 60/70% or samples showing sufficient stability after two days to retrieve near complete genome sequences. For all LFD brands considerable loss of recovery from clinically representative titrations was seen from day three onwards. Interestingly, Martin *et al*., 2022 found that SARS-CoV-2 genomes could be successfully recovered from Abbott and InnoScreen brand test devices up to 8 days after initial sample collection.

The addition of commercial nucleic-acid-stabilising reagents to devices after run completion showed promise. Both RNALater and DNA/RNA shield showed preservative effect over several days upon the typically best yielding devices, OrientGene and Innova, as has been observed by Moso *et al*., 2024 for multiple RNA viruses on the PanBio COVID-19 device. Neither preservative however improved the recovery from the FlowFlex device.

LFDs are not designed to be compatible with extracting genetic material suitable for sequencing. However, building on the observations of *Martin et al*., 2022 and others, we have verified that it is feasible to retrieve adequate genetic material for targeted amplicon sequencing from some of these devices. Our work extends this finding to various LFDs currently deployed in the UK, showing that genomic recovery varies across different devices. Accordingly, a comprehensive assessment of each LFD type is required to determine its suitability as a genomic sequencing sample source.

Consideration should be given as to whether future iterations of LFDs should incorporate the capability to efficiently sequence pathogens from the eluate as a design requirement or how this could otherwise be achieved efficiently and effectively.

We suggest the following next steps:

1. Conduct a comprehensive comparative study to quantify the efficiency of genomic recovery across different LFD brands and models working with representative suppliers. This study should aim to identify factors that influence the variability in recovery rates.
2. Engage with LFD manufacturers to share findings and collaborate on the development of next-generation devices that include the efficient recovery of genetic material as a key design objective.
3. Work towards establishing a set of guidelines or standards for evaluating LFD suitability as sources of genetic material for sequencing. These guidelines may help streamline the assessment process for new or existing LFD types.

## Data Availability

All data produced in the present study are available upon reasonable request to the authors

## Acknowledgements

This study was funded in part by WHO grant 2023/1385507. The authors would like to thank the Porton Diagnostic Evaluation and Innovation team and the wider UKHSA Pathogen Genomics Sequencing Laboratory team for their help and support.

The views expressed are those of the authors and not necessarily those of the funding bodies or the Department of Health and Social Care.

## Supplemental

**Supplementary Table 1.**
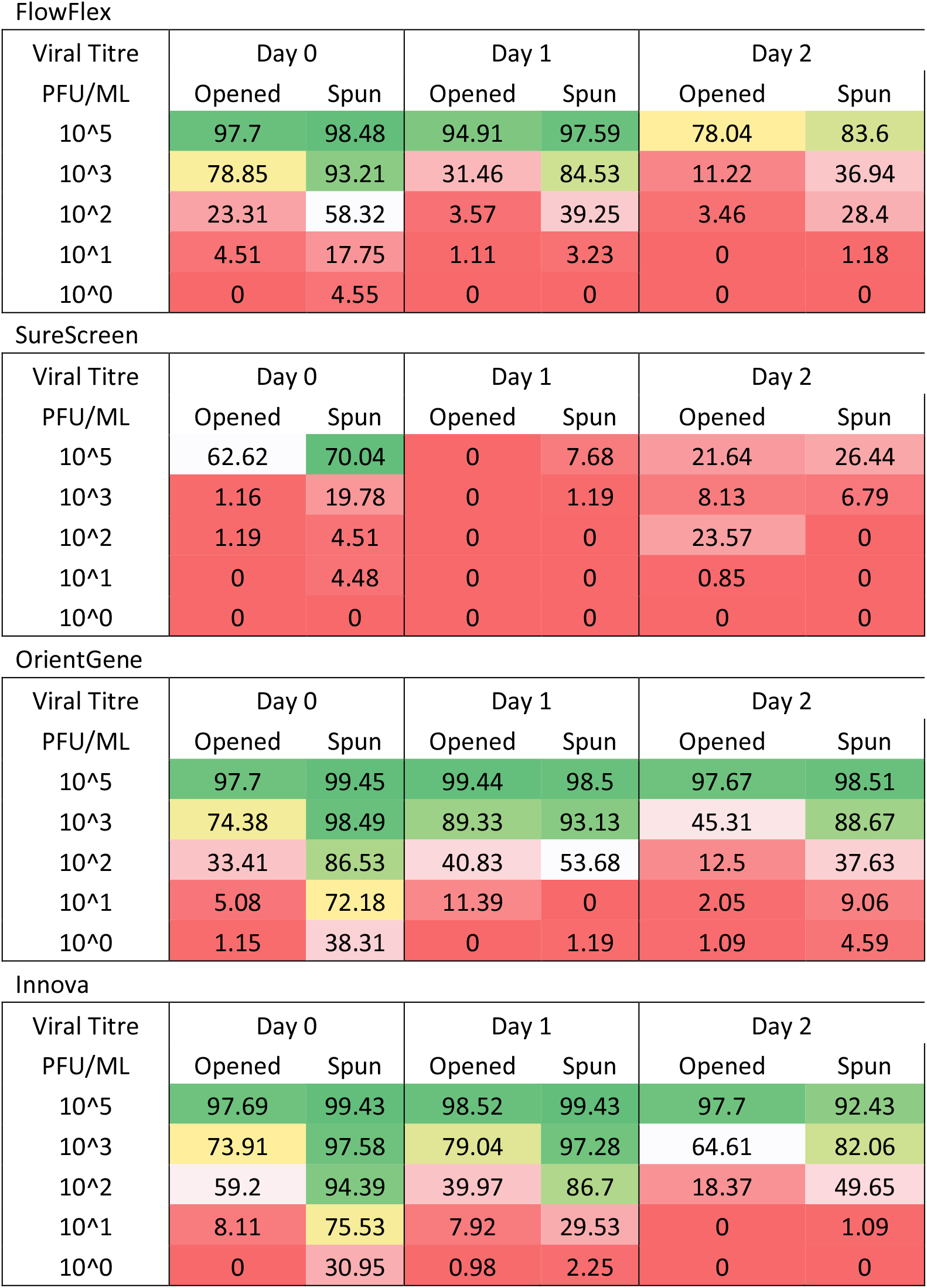
Comparison of percentage consensus genome sequence recovery following elution via strip removal (opened) or centrifugation (spun) method of a range of titres of SARS-COV-2 virions of all four LFD devices tested. Colour coded from green (high) to red (low) coverage.

| Viral Titre<br>PFU/ML | Day 0 |  | Day 1 |  | Day 2 |  |
| --- | --- | --- | --- | --- | --- | --- |
|  | Opened | Spun | Opened | Spun | Opened | Spun |
| 10<sup>5</sup> | 97.7 | 98.48 | 94.91 | 97.59 | 78.04 | 83.6 |
| 10<sup>3</sup> | 78.85 | 93.21 | 31.46 | 84.53 | 11.22 | 36.94 |
| 10<sup>2</sup> | 23.31 | 58.32 | 3.57 | 39.25 | 3.46 | 28.4 |
| 10<sup>1</sup> | 4.51 | 17.75 | 1.11 | 3.23 | 0 | 1.18 |
| 10<sup>0</sup> | 0 | 4.55 | 0 | 0 | 0 | 0 |

SureScreen
| Viral Titre<br>PFU/ML | Day 0 |  | Day 1 |  | Day 2 |  |
| --- | --- | --- | --- | --- | --- | --- |
|  | Opened | Spun | Opened | Spun | Opened | Spun |
| 10<sup>5</sup> | 62.62 | 70.04 | 0 | 7.68 | 21.64 | 26.44 |
| 10<sup>3</sup> | 1.16 | 19.78 | 0 | 1.19 | 8.13 | 6.79 |
| 10<sup>2</sup> | 1.19 | 4.51 | 0 | 0 | 23.57 | 0 |
| 10<sup>1</sup> | 0 | 4.48 | 0 | 0 | 0.85 | 0 |
| 10<sup>0</sup> | 0 | 0 | 0 | 0 | 0 | 0 |

OrientGene
| Viral Titre<br>PFU/ML | Day 0 |  | Day 1 |  | Day 2 |  |
| --- | --- | --- | --- | --- | --- | --- |
|  | Opened | Spun | Opened | Spun | Opened | Spun |
| 10<sup>5</sup> | 97.7 | 99.45 | 99.44 | 98.5 | 97.67 | 98.51 |
| 10<sup>3</sup> | 74.38 | 98.49 | 89.33 | 93.13 | 45.31 | 88.67 |
| 10<sup>2</sup> | 33.41 | 86.53 | 40.83 | 53.68 | 12.5 | 37.63 |
| 10<sup>1</sup> | 5.08 | 72.18 | 11.39 | 0 | 2.05 | 9.06 |
| 10<sup>0</sup> | 1.15 | 38.31 | 0 | 1.19 | 1.09 | 4.59 |

**Supplementary Table 1. Comparison of percentage consensus genome sequence recovery** 370 **following elution via strip removal (opened) or centrifugation (spun) method of a range of** 371 **titres of SARS-COV-2 virions of all four LFD devices tested.**
| Viral Titre<br>PFU/ML | Day 0 |  | Day 1 |  | Day 2 |  |
| --- | --- | --- | --- | --- | --- | --- |
|  | Opened | Spun | Opened | Spun | Opened | Spun |
| 10<sup>5</sup> | 97.69 | 99.43 | 98.52 | 99.43 | 97.7 | 92.43 |
| 10<sup>3</sup> | 73.91 | 97.58 | 79.04 | 97.28 | 64.61 | 82.06 |
| 10<sup>2</sup> | 59.2 | 94.39 | 39.97 | 86.7 | 18.37 | 49.65 |
| 10<sup>1</sup> | 8.11 | 75.53 | 7.92 | 29.53 | 0 | 1.09 |
| 10<sup>0</sup> | 0 | 30.95 | 0.98 | 2.25 | 0 | 0 |

